# Burnout in radiation therapists in Portugal

**DOI:** 10.1101/2020.06.06.20124206

**Authors:** Jéssica Guerra, Francisco Caramelo, Miguel Patrício

## Abstract

**Background:** Burnout syndrome has adverse consequences for individuals, causing a variety of cognitive, affective, physical, behavioural and motivational problems. We aim to assess burnout in Portuguese radiation therapists, for who high levels of contact with patients may potentially lead to burnout.

**Methods:** Radiation therapists working in Portugal were invited via e-mail to participate in the study by filling in a survey. The latter had two components: a social-demographic questionnaire and the Maslach Burnout Inventory – Human Services Survey.

**Results:** A total of 103 people responded to the survey, 95 of which employed as radiation therapists. The mean burnout scores were 20.60 ± 11.21, 7.43 ± 5.34 and 35.02 ± 6.02, for the emotional exhaustion, depersonalisation and personal accomplishment subscales, respectively. In the same order, the total of radiation therapists presenting high levels of burnout were 29%, 14.9% and 29.3% for the different dimensions. The mean scores of burnout did not differ significantly regarding gender, civil status, working in the public or private sector and years of service. Radiation therapists aged 40 years or older presented greater scores of burnout, though with no statistical significance.

**Conclusion:** Radiation therapists working in Portugal were seen to have average scores of burnout in the emotional exhaustion, depersonalisation and personal accomplishment subscales.

## Introduction

Burnout has been an object of study for different health occupations, notably for physicians (1), medicals students (2) and other health care professionals (3). As a construct, burnout must be distinguished from occupational stress, which may theoretically lead to burnout (4). In particular for oncology care, a number of studies have assessed burnout on doctors or nurses, see for example (5–8) and (8–10), respectively. However, few articles have included an evaluation of burnout in radiation therapists (RTs), a workforce that works on a daily basis with cancer patients, high therapeutic doses, heavy workloads and inappropriate support (11).

Some articles do include RTs in their samples (5,12–14) but present their burnout scores pooled with those of other occupations, while other publications focus solely on RTs or explicitly report burnout scores for this population (4,15–23).

The instrument most used to assess burnout is the Maslach Burnout Inventory (MBI) questionnaire, which includes three subscales: emotional exhaustion (EE), depersonalisation (DP) and personal accomplishment (PA) (24,25). According to the authors of the original version of the MBI questionnaire, the MBI - Human Services Survey (MBI-HSS), the three dimensions of the questionnaire measure “feelings of being emotionally overextended and exhausted by one’s work”, “unfeeling and impersonal response toward [patients]” and “feelings of competence and successful achievement in one’s work”, respectively (26).

Few authors have used other questionnaires - notably the Oldenburg Burnout Inventory (27) and the Pro-Quality of Life questionnaire (22) - to assess burnout in RTs, while most have resorted to MBI-HSS (4,15–21). Focusing solely on the latter, the majority of studies surveyed RTs located in North America (16,18,19,21) or in Europe (4,17,28), with a single study from Australia (15). Concerning the results that were reported, regardless of cultural differences, RTs were consistently seen to have average scores of burnout in the three subscales of the MBI questionnaire (4,15,19,20), with exception of Koo, *et al* (16) and Demirci, *et al* (17), that observed low levels of burnout in the DP subscale. In addition, Sale, *et al* (18) reported high levels of burnout in the DP subscale while Akroyd *et al* (21) found that RTs from his USA sample had higher scores of burnout in the EE and DP subscales. Interestedly, this study reports the lowest burnout scores in the PA subscale.

No studies aiming at evaluating the burnout levels of RTs working in Portugal have previously been reported. We aim at assessing burnout in this population using the MBI-HSS questionnaire.

## Material and Methods

### Study design and sample

Radiotherapists residing in Portugal were surveyed in February 2017. The web-based survey, written in Portuguese, had an estimated time of response of 5 minutes and comprised socio-demographic questions (sex, age, marital status, years of service as an RT and whether the participant worked in the public or in the private sector) and the MBI -Human Services Survey (MBI-HSS). The Portuguese association of RTs (ART – Associação de Técnicos de Radioterapia) was contacted to distribute the survey among their associates. Participation was voluntary and the confidentiality of the participants was guaranteed at all stages of the process. Solely the authors of the study had access to the data. No compensation or other reward was provided for participation. The study was reviewed and approved by the Ethics Commission of the Faculty of Medicine of the University of Coimbra.

### Burnout questionnaire

In addition to questions to gather socio-demographic data, participants were asked to respond to the MBI-HSS questionnaire. This questionnaire is composed by 22 items and it is divided in 3 subscales: EE (comprising by 9 items), DP (comprising 5 items) and PA (comprising 8 items). A 7-point Likert scale is used for each item and scores within individual burnout domains can either be used as continuous variables or categorized into indicators of low, average or high levels of burnout using established cut-offs, see Table 1 (29).

**Table 1.** Burnout Scores

| **MBI Subscale** | **Burnout Level** |  |  |
| --- | --- | --- | --- |
|  | Low | Average | High |
| **EE** | $\leq 16$ | 17-26 | $\geq 27$ |
| **DP** | $\leq 6$ | 7-12 | $\geq 13$ |
| **PA** | $\geq 39$ | 38-32 | $\leq 31$ |

It is worthwhile noting that while high levels in the EE and DP subscales are associated to high burnout, high levels in PA subscale are associated to low burnout.

### Statistical Analysis

Categorical data are described by absolute and relative frequencies whereas for quantitative data the means and standard deviations were used instead. The median, 25^th^ percentile, 75^th^ percentile, minimum and maximum of the MBI scores in the three dimensions are also presented. The normality of quantitative variables was checked with Shapiro Wilk tests. Associations between pairs of categorical data and correlations between pairs of quantitative variables were assessed using Fisher tests and by computing Spearman’s correlation coefficients, respectively. Whenever normality assumptions held, t-Student tests and ANOVA were used to verify whether statistically significant differences arose between two groups or more groups, respectively. When the assumptions did not hold, Mann-Whitney and Kruskal-Wallis tests were used instead. The significance level adopted was α=0.05. The statistical analysis was performed on IBM^©^ SPSS^©^ Statistics 24.

## Results

### Participation and sample

A total of 103 people responded to the survey. Eight of these were not employed as RTs at the time. The results below pertain to the remaining 95 (92.2%) participants, 81 of which were female (85.3%) and 14 (14.7%) male, see Table 2. Two of the respondents did not provide information on their age and out of the other 93 that did, the average age was observed to be 30.2±5.9. When asked about their civil status, 53 (55.8%) of the RTs stated they were single, 41 (43.2%) married and one RT divorced (1.1%). Fewer participants – 43 (45.3%) – were working in the public sector than in private RT departments – 52 (54.7). The mean number of years working as an RT was seen to be 7.06 ± 5.59.

**Table 2.** Demographical characterisation

|  | **Subgroup** | **n (%)** |
| --- | --- | --- |
| **Gender** | Female | 81 (85.3%) |
|  | Male | 14 (14.7%) |
| **Age (years)** | [20-29] | 52 (55.9%) |
|  | [30-39] | 36 (38.7%) |
|  | [40-49] | 3 (3.2%) |
|  | 50+ | 2 (2.2%) |
| **Civil Status** | Single | 53 (55.8%) |
|  | Married | 41 (43.2%) |
|  | Divorced | 1 (1.1%) |
| **Sector** | Public | 43 (45.3%) |
|  | Private | 52 (54.7%) |
| **Years in RT service** | [0-4] | 38 (40%) |
|  | [5-9] | 28 (29.5%) |
|  | [10-14] | 21 (22.1%) |
|  | [15-19] | 5 (5.3%) |
|  | 20+ | 3 (3.2%) |

Data are presented as number (percentage).

### Burnout results

The results of the MBI questionnaire are shown in Table 3. Not all participants replied to every item of the questionnaire; the number of responders ranged from 92 for PA to 94 for DP. For EE the mean was seen to be 20.60 ± 11.21; for DP it was 7.43±5.34 and for PA the mean was 35.02±6.02.

**Table 3.** Burnout scores for each of the three dimensions of the MBI-HSS questionnaire.

| **MBI Subscale\*** | **M ± SD [min-max]** | **Med (P<sub>25</sub>; P<sub>75</sub>)** |
| --- | --- | --- |
| **EE (n=93)** | 20.60±11.21 [1-50] | 20 (11; 28) |
| **DP (n=94)** | 7.43±5.34 [0-28] | 7 (3; 11) |
| **PA (n=92)** | 35.02±6.02 [20-48] | 35 (31; 40) |

\*For each MBI subscale, data are presented as mean±standard deviation [minimum-maximum] and as median (25<sup>th</sup> percentile; 75<sup>th</sup> percentile)

As aforementioned, for each subscale, the burnout score can be categorised as low, average or high level. Table 4 displays the percentage of RTs at different levels of burnout in each subscale. For the EE subscale, 36 (38.7%) of the RTs were seen to have low scores, 30 (32.3%) with average scores and 27 (29.0%) with high burnout scores. Better results were found for the DP subscale, as only 14 (14.9%) of the RTs had high scores of burnout, with 46 (48.9%) and 34 (36.2%) with low and average scores of burnout, respectively. For PA, 27 (29.3%) of the RTs were seen to be with high scores, with 35 (38.0%) and 30 (32.6%) being with average and low levels, respectively.

**Table 4.** Prevalence of low, average and high scores of burnout for each of the three dimensions of the MBI-HSS questionnaire.

|  | **Burnout Level** |  |  |
| --- | --- | --- | --- |
|  | Low | Average | High |
| **EE (n=93)** | 36 (38.7%) | 30 (32.3%) | 27 (29.0%) |
| **DP (n=94)** | 46 (48.9%) | 34 (36.2%) | 14 (14.9%) |
| **PA\* (n=92)** | 30 (32.6%) | 35 (38.0%) | 27 (29.3%) |

Data are presented as number (percentage).

### Relationship between burnout components, demographics and work history

When the average EE, DP and PA scores are considered, RTs were seen to nearly always be with average scores of burnout, independently of gender, age, civil status, working in the public or private sector and years of service, see Table 5. More detailed information about the numbers of individuals at each level of burnout can be found on Supplementary material. There were few exceptions where the mean MBI scores where not indicative of average levels of burnout. Notably, for RTs aged between 20 and 29 years old and RTs working in the area for up to 4 years presented low levels of burnout in the DP subscale; RTs with 40 years or more presented high levels of burnout in the EE and PA subscale.

**Table 5.** Burnout scores for different subgroups.

|  |  | EE | *p*-value | DP | *p*-value | PA | *p*-value |
| --- | --- | --- | --- | --- | --- | --- | --- |
| **Gender** | Female | 19.8±10.<br>8 | 0.160 | 7.0±5.0 | 0.106 | 35.2±6.<br>0 | 0.464 |
|  | Male | 25.3±12.<br>9 |  | 10.0±6.<br>6 |  | 33.9±6.<br>2 |  |
| **A g e**<br>**groups** | [20-29] | 19.0±10.<br>8 | 0.199 | 6.6±4.5 | 0.500 | 35.8±5.<br>7 | 0.192 |
|  | [30-39] | 20.9±10.<br>8 |  | 8.2±6.0 |  | 34.7±6.<br>4 |  |
|  | 40+ | **29.2±15.**<br>**2** |  | 8.4±7.4 |  | **30.8±5.**<br>**3** |  |
| **C i v i l**<br>**Status** | S i n g l e /<br>divorced | 21.0±11.<br>3 | 0.716 | 7.6±4.6 | 0.766 | 34.6±5.<br>6 | 0.424 |
|  | Married | 20.1±11.<br>2 |  | 7.1±6.2 |  | 35.6±6.<br>6 |  |
| **Sector** | Public | 22.3±11.<br>8 | 0.147 | 7.9±5.0 | 0.271 | 34.9±6.<br>0 | 0.812 |
|  | Private | 19.2±10.<br>6 |  | 7.0±5.6 |  | 35.2±6.<br>1 |  |
| **Years of**<br>**service**<br>**a s a n**<br>**RT** | [0-4] | 17.2±10.<br>8 | 0.092 | 6.8±4.4 | 0.641 | 35.6±5.<br>9 | 0.634 |
|  | [5-9] | 22.5±11.<br>5 |  | 7.4±6.7 |  | 34.9±6.<br>1 |  |
|  | [10-14] | 21.7±9.6 |  | 8.3±4.6 |  | 35.2±6.<br>8 |  |
|  | 15+ | 26.4±13.<br>5 |  | 8.0±6.6 |  | 32.5±4.<br>6 |  |

\* For each of the three dimensions (EE, DP and PA) and for each category of each variable, data are presented as mean±standard deviation. Results presented in cells with grey background correspond to lower levels of burnout (normal font) or higher levels of burnout (boldface).

Females shown slightly lower levels of burnout in the EE and PA subscales than males, although no statistically significance was found in either subscales (*p*=0.160 and *p*=0.106, respectively). Better results were also found for females in the PA subscale (*p*=0.464).

When analysing whether the age groups of the RTs were associated with the burnout scores, it was found that the median burnout scores to EE, DP and PA were similar, independently of the group *p*=0.199, *p*=0.500 and *p*=0.192, respectively), except for RTs with 40 years or more, who presented high levels of burnout in the EE and PA subscales. As for RTs aged between 20 and 29, they had low burnout scores in the EE subscale. Correlations between age and the values obtained in the three dimensions were further assessed. A statistically significant weak positive correlation was found between age and EE (r_S_=0.219, p=0.037), but no significant correlations were attained between age and DP (r_S_=0.030, p=0.778) or age and PA (r_S_=-0.044, p=0.682).

Regarding the marital status of the participants, the median burnout score for each subscale was similar between RTs who were single or divorced and RTs who were married (*p*=0.716, *p*=0.766 and *p*=0.424, respectively for the EE, DP and PA subscales). People working in the public sector had EE and DP values higher than the ones observed in RTs working in the private sector; however this difference was not statistically significant (*p*=0.147 and *p*=0.271, respectively). In the PA subscale, the results showed that RTs working in the public sector were at slightly higher level of developing burnout (*p*=0.812).

No statistically significant associations were found between years in service as an RT and burnout levels (*p*=0.092, *p*=0.641 and *p*=0.634, respectively for the EE, DP and PA subscales). However, RTs with less years of services displayed better levels of burnout in the three subscales.

## Discussion

Burnout syndrome has adverse consequences for individuals, causing a variety of cognitive, affective, physical, behavioural and motivational problems, (30). This syndrome can decrease the quality of life of workers and often induces the increase of anxiety, depression, headaches, mood swings, hypertension and cardiovascular or gastrointestinal disorders. Job performance, dissatisfaction, absenteeism, intention to leave the job, low levels of commitment to the organisation, lower productivity or morale and turnover are other facets that have been frequently associated to burnout. (7,24–26).

RTs practice a caring profession known to often have high levels of contact with patients, which may potentially lead to burnout (4). In our study, the burnout scores of the RT population working in Portugal was seen to be 20.60 ± 11.21, 7.43 ± 5.34 and 35.02 ± 6.02 for the EE, DP and PA subscales, respectively, all translating an average burnout score. In this sense, the results were similar to those reported in the literature. However, for PA, the average score observed was worse than those reported by every other study assessing burnout in RTs. In the Portuguese RTs, PA was 35.02 ± 6.02, while for other countries the average PA ranged between 36.0 (± 6.8) and 42.1 (± 6.3), see (18) and (21), respectively.

The percentage of RTs with high levels of burnout in Portugal was seen to be 29.0%, 14.9% and 29.3% for the EE, DP and PA subscales, respectively. Focusing on EE, for which the literature reports high burnout scores to range between 19.5% and 55%, only two authors observed lower burnout percentages (15,16). The proportion of RTs at high burnout scores in the DP dimension ranges from 1.8% to 45% in the literature, with only Diggens et al. (15) reporting a lower proportion than what was observed for the Portuguese sample. For PA, the literature reports high scores percentages from 16.8% up to 58.3%, except Koo K et al. (16) that exceeded, quite expressively, what was seen for the Portuguese RTs.

Several authors studied potential associations between burnout and individual or workplace factors, not specifically for RTs. In particular, younger workers are seen to normally present higher levels of burnout than those 30 years or older; gender has not been shown to be strongly associated to burnout; unmarried workers seem to have higher burnout levels than those who are married or divorced; burnout seems to be higher in workers with a higher education level, (25). As “burnout is more of a social phenomenon than an individual one”; factors such as workload, control, reward, community, fairness and values may be more relevant (24,25). For RTs, relationships were found to hold between burnout and age, marital status and number of years of experience (17) or between burnout, age and gender (18). In the current study, an analysis of the relationship between social-demographic and work history of the RTs and the three burnout subscales was performed. Older RTs (≥ 40 years) were seen to have higher levels of burnout in all subscales, though no statistical significance was reached.

Response bias is a potential limitation of this study, as it is unknown whether professional and personal distress weighed on the response rate. Furthermore, it should be noted that to avoid discouraging potential responders from filling out the survey that was conducted for this study, only a small number of questions were included in it besides the MBI-HSS questionnaire. Though this is not a large study, the sample size is quite considerable given the number of RTs in Portugal. However, this attempt to maximise the number of responders came at the cost of exploring possible causes or correlates of burnout in Portuguese RTs. Adding questions on job satisfaction, job characteristics or even additionally resorting to quality of life or stress surveys could have been informative. In conclusion, our results indicate a prevalence of burnout in RTs in Portugal which is similar to what is reported in the literature, with RTs aged 40 years or more presenting greater burnout scores.

## Conclusion

In our study we observed that RTs in Portugal have average burnout levels in the EE, DP and PA subscales, evaluated by the MBI-HSS questionnaire, with 29%, 14.9% and 29.3% being with high burnout scores, respectively. These results are similar to what had been reported in the literature for other countries.

## Data Availability

The data that support the findings of this study are available from the corresponding author, [MP], upon reasonable request.

## Acknowledgements

Research was conducted in CNC.IBILI and supported by the Foundation for Science and Technology (Ref. UID/NEU/04539/2013) and COMPETE 2020 - Operational Program for Competitiveness and Internationalization (Ref. POCI-01-0145-FEDER-007440). The authors thank ART (Associação de Técnicos de Radioterapia) for the online distribution of the questionnaires.

## Supplementary Material

**Table 6.** Prevalence of low, average and high burnout scores for each of the three dimensions of the MBI-HSS questionnaire, for different subgroups.

|  |  | EE | *p*-<br>valu<br>e | DP | *p*-<br>valu<br>e | PA | *p*-<br>valu<br>e |
| --- | --- | --- | --- | --- | --- | --- | --- |
| **Gender** | Female (n=81) | 41% :<br>33% : 27% | 0.50<br>3 | 53% :<br>35% : 13% | 0.13<br>9 | 32% :<br>41% : 27% | 0.31<br>4 |
|  | Male (n=14) | 29% :<br>29% : 43% |  | 29% :<br>43% : 29% |  | 36% :<br>21% : 46% |  |
| **Age**<br>**groups** | [20-29] (n=52) | 46% :<br>32% : 22% | 0.42<br>0 | 51% :<br>40% : 10% | 0.44<br>4 | 41% :<br>35% : 25% | 0.27<br>8 |
|  | [30-39] (n=36) | 33% :<br>36% : 31% |  | 47% :<br>36% : 17% |  | 28% :<br>52% : 31% |  |
|  | 40+ (n=5) | 20% :<br>20% : 60% |  | 40% :<br>20% : 40% |  | 0% : 40% :<br>60% |  |
| **Civil**<br>**Status** | Single or<br>divorced (n=54) | 37% :<br>35% : 29% | 0.83<br>7 | 45% :<br>42% : 13% | 0.47<br>7 | 31% :<br>37% : 31% | 0.89<br>8 |
|  | Married (n=41) | 42% :<br>29% : 29% |  | 54% :<br>29% : 17% |  | 34% :<br>39% : 27% |  |
| **Sector** | Public (n=43) | 33% :<br>29% : 38% | 0.23<br>0 | 42% :<br>40% : 19% | 0.42<br>0 | 33% :<br>33% : 35% | 0.52<br>0 |
|  | Private (n=52) | 43% :<br>35% : 22% |  | 55% :<br>33% : 12% |  | 33% :<br>42% : 25% |  |
| **Years of**<br>**service as**<br>**an RT** | [0-4] (n=38) | 53% :<br>28% : 19% | 0.40<br>4 | 49% :<br>43% : 8% | 0.20<br>9 | 39% :<br>39% : 22% | 0.55<br>9 |
|  | [5-9] (n=28) | 32% :<br>36% : 32% |  | 57% :<br>25% : 18% |  | 37% :<br>30% : 33% |  |
|  | [10-14] (n=21) | 29% :<br>38% : 33% |  | 38% :<br>48% : 14% |  | 24% :<br>48% : 29% |  |
|  | 15+ (n=8) | 25% :<br>25% : 50% |  | 50% :<br>13% : 38% |  | 33% :<br>38% : 50% |  |

For each of the three dimensions (EE, DP and PA) and for each category of each variable, data is presented as percentage of people with low burnout levels : percentage of people with average burnout levels : percentage of people with high burnout levels; n=sample size.

